# Short-Chain Oat Fiber Improves Gastrointestinal Tolerance and Regulates Glucose Metabolism: A Two-Week Open-Label Study in Healthy Adults

**DOI:** 10.64898/2026.01.21.26343795

**Authors:** Angela M Marcobal, Katharine M Ng, Riley A Drexler, Bruce R McConnell, Matthew J Amicucci

**Author notes:** **Correspondence:** Matthew J. Amicucci.

## Abstract

**Introduction:** Fiber intake is the most common nutritional inadequacy in the Western diet, with most adults consuming less than half of the recommended intake with only 5% of adults meeting the RDI. A novel, short-chain beta-glucan derived from oats (scOat Fiber), with improved solubility, low viscosity and enhanced palatability, compared to conventional oat fibers, was investigated for its benefits as a source of fiber supplementation.

**Methods:** **A** 14-day pilot study evaluated the gastrointestinal tolerance and functional benefits of scOat Fiber in 63 healthy adults randomized to receive 5, 10 or 20 g daily doses. The primary outcome, gastrointestinal tolerability, was assessed using the Gastrointestinal Symptom Rating Scale (GSRS). Secondary outcome included glycemic response during rice challenges, measured via continuous glucose monitoring (CGM). CGM was also used to explore overall glucose dynamics. Additional exploratory outcomes (mood, energy, appetite and sleep) were assessed via validated questionnaires.

**Results:** scOat Fiber was exceptionally well tolerated across all doses, with no increase in GSRS scores, which remained in the low to mild range. Significant reductions in total GSRS scores were observed, with benefits evident after just one week at 5 g/day and maintained over time at both 5 and 10 g/day groups. Evaluation of GSRS sub-categories revealed that the 5 g/day and 10 g/day dose groups experienced significant reductions in abdominal pain symptoms. Both dose groups also demonstrated a significant decrease in constipation at the end of the study. Postprandial glucose responses were attenuated following product use, with a significant reduction in peak glucose during rice challenges after 2 weeks in the 20 g/day group. Both 10 and 20 g/day doses were associated with significant improvement in glycemic metrics during the study, including reductions in glucose mean, all glycemic excursions, and an increase in time-in-range. Exploratory analysis suggested that scOat Fiber may improve mental health and concentration in participants with elevated baseline symptoms.

**Conclusions:** Despite the lack of a placebo control and short duration, the dose-dependent nature of the results supports the potential of scOat Fiber as a well-tolerated and functional source of fiber with benefits including glycemic control, digestive health and mental health (NCT06739941)

## 1 Introduction

Dietary fiber plays a key role in contributing to and maintaining human health through various pathways such as modulating glycemic response, cholesterol levels, blood pressure, mineral absorption, gut barrier function and the gut microbiota (Eswaran et al., 2013; Scholz-Ahrens et al., 2007; Slavin, 2013). Conversely, a lack of dietary fiber intake is associated with many chronic conditions (McRae, 2018, 2017; Timm et al., 2013). However, according to the 2020-2025 *Dietary Guidelines for Americans*, only 10% of women and 3% of men, consume the recommended daily fiber intake, and in general, the adult population in the US consumes less than 50% of adequate fiber amounts (Storz and Ronco, 2022). This fiber gap is further exacerbated by growing popularity of low carbohydrate diets, such as the ketogenic, paleo or carnivore, which often restrict or eliminate fiber-rich plants, including whole grains in favor of animal-based proteins and fats (Storz and Ronco, 2022).

With 340 million people in the United States running a 16 g/day gap in dietary fiber intake, this presents a 2 billion kilogram opportunity for the food, beverage and supplement industries to develop fiber enriched products that can dramatically improve public health through the reduction of chronic disease and all-cause mortality (Reynolds et al., 2019). However, challenges in formulation and gastrointestinal tolerance issues have limited the use of natural dietary fibers in ready-to-eat and drink food products. Most dietary fibers isolated from natural plant sources or food industry side streams have physical characteristics that complicate their formulation, such as gritty texture and tendency to gel, leading to undesirable changes in organoleptic and sensory properties (De Santis et al., 2020). As a result, modified or synthetic fibers, such as corn fiber, wheat dextrin or cassava root fiber, have been increasingly used in foods and supplements, despite their limited benefits (Holscher, 2017). Inulin, commonly extracted from chicory root, agave, or Jerusalem artichoke, is a popular alternative that is more compatible with the formulation needs of the food industry. However, when formulated into food and beverages with acidic pH (pH<5), inulin is chemically fragile, breaking down into fructose and consequently increasing sugar content (Martins et al., 2019). In addition, the tolerability of inulin is a concern. Five to ten g/day doses of inulin are required to achieve health benefits, but they are frequently associated with gastrointestinal side effects, including gas, bloating, and abdominal pain (Bruhwyler et al., 2009; Holscher et al., 2022). Also, recent studies have associated the consumption of high concentrations of inulin with inflammation in sensitive individuals (Arifuzzaman et al., 2024, 2022).

Mixed linkage β-glucans, are polysaccharides naturally present in several grains such as oat, barley, rye and wheat (Lazaridou and Biliaderis, 2007) and present an opportunity for fiber supplementation. As human digestive enzymes are unable to cleave the β1-4 and β1-3 glucose linear bonds of β-glucans, these structures pass into the colon where they are metabolized by the gut microbiota (Tamura et al., 2017). Cereal β-glucans have been acknowledged by regulatory bodies for their positive impact on cardiovascular and metabolic health [13-15]. However, oat β-glucans are extremely gelling, and their thick mouthfeel makes them unsuitable for many applications, especially beverages. β-glucans are also stable over most conditions used in food and beverages (Ren et al., 2018).

To exploit the benefits of oat fiber while mitigating its formulation issues, a novel, short-chain oat fiber product (scOat Fiber), obtained by depolymerization of natural oat β-glucan, has been developed. Traditionally, the physiological benefits of long-chain oat β-glucans improving blood glucose profiles are attributed to their high viscosity, which delays gastric emptying and slows nutrient absorption in the small intestine. In contrast, the depolymerized short-chain form exhibits minimal viscosity yet retains distinctive functional properties. It has recently been shown to exert a prebiotic effect on human microbial communities, with robust production of short chain fatty acids (SCFA) (Maldonado-Gomez et al., 2025). *In vitro* fecal fermentations using multiple human stools, revealed an outstanding fermentability profile and high production of beneficial SCFAs, including butyrate. Additionally, the impact of this novel fiber product on mechanisms related to glucose control, such as inhibition of key digestive enzymes and sodium glucose co-transporters (SGLT-1), have been demonstrated *in vitro*, suggesting a capacity to slow down carbohydrate digestion and glucose uptake (Marcobal et al., 2024).

Although the gastrointestinal tolerance of scOat Fiber is expected to be similar to native oat fiber, until now, this has not been shown. Further, the potential benefits suggested by preclinical studies have not been validated in humans. The aim of this dose ranging study in healthy adults was to assess the tolerability, potential digestive health and metabolic effects of the novel oat fiber ingredient in a real-world setting. Participants were asked to consume three different doses (5, 10 and 20 g/day), for 2 weeks. The doses were chosen to reflect a range of realistic applications in products, and to assess tolerability at a high daily dose. The high daily dose was at a level that normally exceeds the gastrointestinal (GI) discomfort threshold seen with other fibers, such as inulin or psyllium husk. This study also explored outcomes including mood, energy, appetite, and sleep. The study was also intended to inform the design of a future, large, controlled study.

## 2. Materials and methods

### 2.1 Trial design and conduct

This study was a prospective, open-label, three-arm clinical study to evaluate the gastrointestinal tolerance and safety of daily oral intake of 5g, 10g and 20g doses of scOat Fiber in healthy adults. The scOat Fiber (one.bio Oat Fiber) was manufactured by One Bio Inc. (Sacramento, CA, USA) by depolymerizing oat fiber. The scOat Fiber comprises mainly β-glucan oligosaccharides, with a small amount of arabinoxylan oligosaccharides. Most oligosaccharides have short-chain lengths of 3 to 30 monomer units. The study was conducted by People Science Inc., in accordance with the ethical principles outlined in the Belmont Report: Ethical Principles and Guidelines for the Protection of Human Subjects of Research (US National Commission for the Protection of Human Subjects of Biomedical and Behavioral Research, April 18, 1979) and the Declaration of Helsinki. The study was conducted according to the US Code of Federal Regulations Title 45 and State of California Health and Safety Code. The protocol was approved by the Advarra IRB (Columbia, MD, USA; Protocol ID#Pro00074413) and registered with ClinicalTrials.gov (NCT06739941). All participants were recruited from the community through social media channels and researcher networks and all provided written informed consent prior to starting the study.

After inclusion in the study, the participants completed an informed consent form and provided demographic information, medical history, dietary habits, and physical measurements. Complete Blood Count (CBC), Comprehensive Metabolic Panel (CMP), and hemoglobin A1C tests were performed at a Quest Lab to assess study eligibility. Eligible participants were then randomly assigned to one of three product dose groups (5 g, 10 g, or 20 g of scOat Fiber per day), after which the product was shipped to participants. The study concluded with follow-up blood tests. The participants also completed a standardized at-home rice challenge to evaluate postprandial glucose responses. The challenge involved consuming 210 g (7.4 oz) of ready-to-eat cooked rice (NISHIKI® brand), providing approximately 59 g of available carbohydrate, after an overnight fast of 8–10 hours. The rice was consumed alone prior to commencing scOat Fiber use (intervention T0), and in combination with scOat Fiber at the start of scOat Fiber consumption (intervention 1, T1), after 1 week of daily scOat Fiber consumption (intervention 2, T2), and after two weeks of daily scOat Fiber (intervention 3, T3). An overview is shown in Figure 1. All data were collected and securely stored on a platform (Chloe, Consumer Health Learning & Organizing Ecosystem) run by People Science Inc. on Amazon Web Services HIPAA-compliant servers.

**Figure 1.**
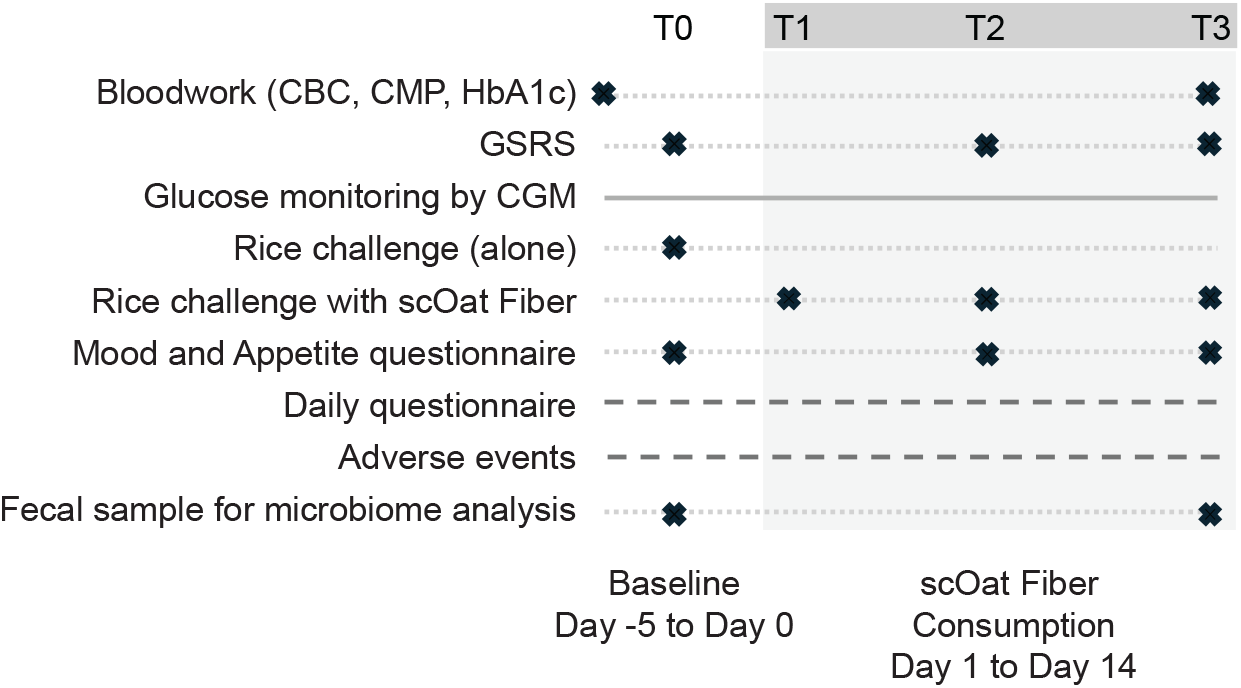
Clinical study design. Data was collected continuously (grey line), daily (dashed line) and at specified timepoints (dotted lines). Intervention times are shown as T0 (rice challenge alone), T1 (at the start of fiber consumption), T2 (after 1 week of daily fiber consumption) and T3 (after 2 weeks of daily fiber consumption). Daily questionnaire includes information related to absence of GI symptoms, unusual meals, unusual drop of energy, sleep quality and time to fall asleep.

The validated Gastrointestinal Symptom Rating Scale (GSRS) was used to evaluate tolerability of scOat Fiber [18]. GSRS scores are grouped into five sub-categories (reflux, abdominal pain, indigestion, diarrhea, and constipation) or summarized as a total score. The GSRS questionnaire was completed at intervention T0, after one week of fiber consumption (intervention T2), and at the end of the study (intervention T3). Participants also completed:

1. A weekly questionnaire with 12 questions assessing appetite, anxiety, depression and concentration. This weekly questionnaire consisted of a survey drawn from custom questions, and a subset of items in the validated Generalized Anxiety Disorder 7-Item (GAD-7) and the Patient Health Questionnaire 9-Item (PHQ-9),
2. A daily questionnaire consisting of five questions that enquired about (i) presence or absence of gastrointestinal (GI) symptoms, (ii) whether an unusual meal was consumed (to potentially explain unusual symptoms, glucose peaks), (iii) unusual drops in energy, (iv) sleep quality, and (v) time taken to fall asleep.
3. A daily adverse event questionnaire to report on any negative effects they may have experienced during product consumption.

Participants also wore a Dexcom G6 Pro Continuous Glucose Monitoring (CGM) device for 19 days (baseline period and intervention period) to evaluate blood glucose dynamics using multiple metrics.

### 2.2 Inclusion and exclusion criteria

Participants were eligible for participation in the study if they were aged between 18 years and 70 years, were able to read and understand English, understand and provide informed consent, use a personal smartphone device and to complete study assessments over the course of up to 7 weeks. Participants were excluded if they did not have a smartphone and/or internet access, had any significant abnormal laboratory value seen on CBC and CMP testing and/or an A1C > 6.5%, were diagnosed with a gastrointestinal, digestive or metabolic disease or disorder including Crohn’s disease, ulcerative colitis, irritable bowel syndrome, celiac disease, gluten allergy, diabetes, or obesity, or an illness, disease or condition which, in the opinion of the principal investigator, may have impacted their ability to participate in the study or impacted the study outcomes, had any known allergic reaction to any component of the scOat Fiber or the rice challenge product, were prescribed medication likely to influence the study measures, had consumed fiber supplements 30 days prior to enrollment or commenced consumption at any time during the study, had undergone a major change in diet or exercise 30 days prior to enrollment or who initiated such a change at any time during the study, had consumed antibiotics 30 days prior to enrollment and who initiated such consumption at any time during the study, were pregnant, wished to become pregnant, or who were breastfeeding, had excessive alcohol use or substance abuse, were unlikely for any reason to be able to comply with the study protocol or who were considered unsuited for participation in the study by the principal investigator.

### 2.3 Study outcomes

The primary outcome was gastrointestinal tolerability of the 3 different doses of scOat Fiber as measured by change in mean total score on the Gastrointestinal Symptom Rating Scale (GSRS) at interventions T2 and T3 with respect to Intervention T0.

Secondary and exploratory outcomes included:

1. The effects on gastrointestinal symptoms of the 3 different doses of scOat Fiber as measured by change in mean total score, and the 5 sub-category scores on the Gastrointestinal Symptom Rating Scale (GSRS) at interventions T2 and T3 with respect to Intervention T0.
2. The effects of a single intervention of the 3 different doses of scOat Fiber on postprandial glucose uptake after a white rice challenge, as measured by iAUC and peak glucose levels using a CGM device at intervention T0 and intervention T1.
3. The effects of two weeks of intervention of the 3 different doses of scOat Fiber on postprandial glucose uptake after a white rice challenge, as measured by iAUC and peak glucose levels using a CGM device at intervention T0 and intervention T3.
4. The effects of the 3 different doses of scOat Fiber on glucose uptake dynamics as measured by a CGM device used during baseline and throughout product use period.
5. The effects of the 3 different doses of scOat Fiber on appetite, anxiety, mood, irritability, anhedonia, concentration and sleep as measured by survey questions.

Safety was assessed based on CBC and CMP test results and monitoring adverse events (AEs) throughout the course of the study.

### 2.4 Statistical analysis

All data analyses were conducted using GraphPad Prism (GraphPad Software, Boston, MA). Normality was assessed using the Shapiro-Wilk (SW) test. Parametric statistics (ANOVA, Student’s t-tests) were used to evaluate differences between normally distributed measures. Non-parametric statistics were used in cases where data were not normally distributed (Friedman’s Test for within-doses across time points, Wilcoxon for post-hoc analyses). The Two-stage step-up method of Benjamini, Krieger and Yekultieli was used to control the False Discovery Rate in the case of multiple comparison testing. Mann-Kendall (MK) tests were used to evaluate potential trends over time in daily data. Fisher’s Exact Test (FET) was used to evaluate potential differences in % improvers.

For the primary outcome, data from all participants who completed intervention T1 were used in the analysis, regardless of whether they completed the study. The same approach was taken for the secondary outcome relating to effects on gastrointestinal symptoms. For the blood glucose related outcomes, participants were considered eligible for analysis if they completed the rice challenge tests at interventions T0 and T3 and had at least 90% CGM data coverage during each rice challenge. Data missing at interventions T1 and T2 were imputed using linear projection. The glucose baseline was defined as the average blood glucose value during the 2 hours fasting period prior to rice consumption. Glucose metrics, including peak glucose, glucose spike height, total area under the curve (AUC), and incremental AUC (iAUC) were calculated using Simpson’s Rule (i.e. trapezoids with parabolic upper edges [19–21]). These metrics were aggregated across participants for each rice challenge test day. The peak glucose was defined as the highest value recorded within 4 hours after rice intake. The glucose spike height was calculated as the difference between the peak glucose and glucose baseline. To evaluate overall impact on glycemic control, the blood glucose data were analyzed longitudinally. The analysis included only participants with complete CGM data across all assessed time points. Metrics were calculated for the baseline period (prior to intervention T1) and for the final five days of product use before Intervention 2 and before intervention 3. Metrics for glycemic control included glucose mean, standard deviation (SD), % coefficient of variation (% CV), excursion size of all excursions, and mean amplitude of glycemic excursions (MAGE). MAGE was calculated per individual on a daily basis as previously described [22,23], then averaged across days and participants. In addition, time in range (TIR) was assessed using both conventional (TIR, 70-140 mg/dL) and a more stringent “ideal” range (iTIR 72-110 mg/dL).

## 3 Results

### 3.1 Sample size and Demographics

67 participants were randomized into three dose groups (5 g, 10 g and 20 g/day), and 63 were evaluated (5 g/day, n=24; 10 g/day, n=19; 20 g/day, n=20). Three participants were withdrawn due to individual participant decisions unrelated to the scOat Fiber, and one participant was withdrawn due to an adverse event related to GI symptoms. Demographics are described in Table 1.

**Table 1.**
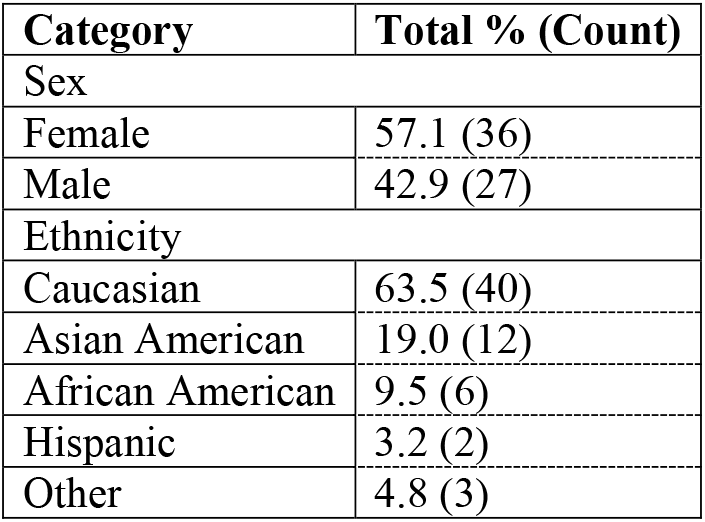
Study demographics.

### 3.2 Safety and tolerance of scOat Fiber

scOat Fiber was well tolerated across all groups, with no increase for any of the three doses groups in total GSRS scores, in any of the 5 GSRS domain scores, and in any of the fifteen GSRS symptom scales, over the course of the study. A difference between the groups was detected (Kruskal-Wallis, *p*= 0.0268), driven by a combination of symptom improvement in two of the groups, and differences in baseline mean scores.

Reported adverse events were predominantly mild and included gas (n=26 participants), bloating (n=15 participants), constipation (n=13 participants), abdominal pain (n=7 participants) and diarrhea (n=5 participants) (Table S2). The number of AEs did not exhibit any dose-dependent relationship. These symptoms were transient, not dose limiting, and none were assessed to be clinically significant. All adverse events were assessed to be possibly related to scOat Fiber or the rice challenge product. One participant discontinued the study due to GI symptoms after intervention 1 (ie, after first consumption of scOat Fiber). The participant was found to be concurrently consuming the antiemetic medication Zofran (Ondansetron). In view of the concurrent use and known side effects of Zofran, no conclusive link between the scOat Fiber and the adverse effect could be established, and the relationship was rated as “Possible”. The participant fully recovered. Analysis of the blood samples revealed no irregularities considered due to the intake of the scOat Fiber in any dose group. All parameters measured remained within the normal range throughout the intervention and any differences that were statistically significant, were not considered to be clinically relevant (data not shown).

Taken together, these data demonstrate that scOat Fiber is safe and well tolerated at daily intakes up to 20 g/day.

### 3.3 Effects on gastrointestinal symptoms

The participants were healthy adults with no or very low gastrointestinal symptoms when entering the study. Despite this, the 5 and 10 g/day dose groups exhibited significant improvement in gastrointestinal symptoms over time, shown by a decrease in total GSRS scores (Table 2, Friedman *p*= 0.0012 and *p*= 0.0105, respectively). Post hoc testing revealed that the 5 g/day group had a significant improvement in Total GSRS score between intervention T0 and interventions T2 and T3 (Table 2, FDR-Adjusted Wilcoxon, *p*= 0.0009, *p*= 0.0026, respectively). Similarly, the 10 g/day group showed improvement in total GSRS between intervention T0 and T3 (Table 2, FDR-Adjusted Wilcoxon, *p*= 0.0124). The total GSRS score for the 10 g/day group at intervention T0 was significantly lower than that of the 5 g/day group (Kruskal-Wallis, *p*= 0.0187), impacting the ability to detect symptom improvement in the 10 g/day group.

**Table 2.**
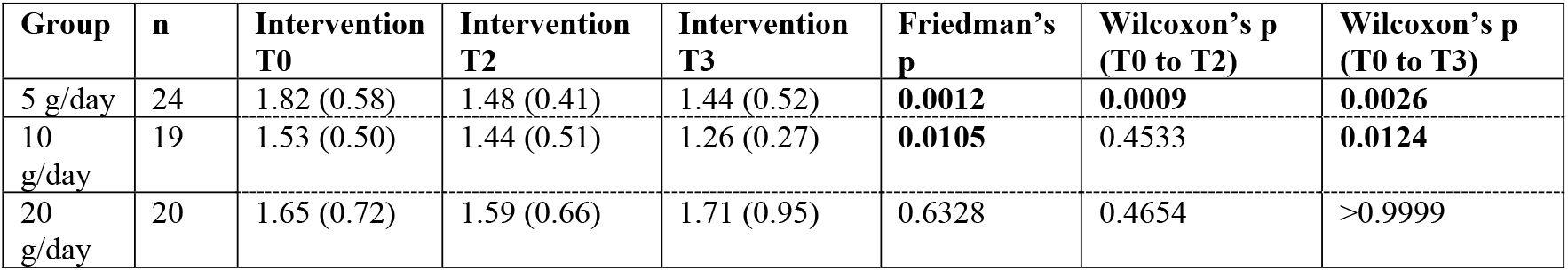
GSRS total scores across the study period. Values represent mean (SD) for each group receiving scOat Fiber at different doses. Statistical significance of changes over time (Friedman’s p value) and between intervention T0 and interventions T2 and T3 (FDR-adjusted Wilcoxon’s p value) are reported as p-values. Significant results (*p*< 0.05) are highlighted in bold.

| Group | n | Intervention T0 | Intervention T2 | Intervention T3 | Friedman's p | Wilcoxon's p (T0 to T2) | Wilcoxon's p (T0 to T3) |
| --- | --- | --- | --- | --- | --- | --- | --- |
| 5 g/day | 24 | 1.82 (0.58) | 1.48 (0.41) | 1.44 (0.52) | **0.0012** | **0.0009** | **0.0026** |
| 10 g/day | 19 | 1.53 (0.50) | 1.44 (0.51) | 1.26 (0.27) | **0.0105** | 0.4533 | **0.0124** |
| 20 g/day | 20 | 1.65 (0.72) | 1.59 (0.66) | 1.71 (0.95) | 0.6328 | 0.4654 | >0.9999 |

When analyzing the GSRS domains, the 5 g/day and 10 g/day dose groups exhibited a significant reduction of abdominal pain symptoms over time (Friedman, *p*= 0.0005 and *p*= 0.0074, respectively, Table S1). Post hoc testing revealed that the 5 g/day group exhibited a significant reduction of abdominal pain symptoms between intervention T0 and interventions T2 and T3 (FDR-adjusted Wilcoxon, *p*= 0.0012 and *p*= 0.0122, respectively). The 10 g/day group experienced a significant reduction of abdominal pain at intervention T3 compared with intervention T0 (FDR-adjusted Wilcoxon, *p*= 0.0187). Additionally, a significant decrease in constipation symptoms was observed over time for the 5 g/day and 10 g/day dose groups (Friedman, *p*= 0.0408 and *p*= 0.0204), and a pairwise significance was found in post-hoc comparisons of intervention T0 and intervention T3 for the 10 g/day groups (Wilcoxon, *p*= 0.0244). No change over time in the other 4 GSRS domain scores was detected, but a significant decrease of indigestion symptoms was found for the 5 g/day dose group when comparing intervention T0 vs intervention T2 (FDR-adjusted Wilcoxon, *p*= 0.0465, Table S1).

Given that bloating and passing gas are common complaints for fibers, these GSRS scales were analyzed separately from the indigestion domain they are part of. No increase in symptoms was detected in any of the groups at any intervention time point. The 5 g/day group in fact exhibited an improvement over time in passing gas symptoms (Friedman, *p*= 0.026).

### 3.4 Impact of scOat Fiber on postprandial glucose

A total of 38 participants were included in the analysis across the three dose groups: 5 g/day (n=13), 10 g/day (n=13), and 20 g/day (n=12). Glucose data from 25 study participants were excluded from the analysis due to (a) missing baseline glucose challenge data (n=6), (b) self-reported failure to fast (n=2), (c) improper reporting of rice challenge timing (n=2), and (d) absence of glucose data (n=15). Blood glucose metrics did not differ between the dose groups at intervention T0 (*p*> 0.05), data not shown.

To understand the dynamics of postprandial blood glucose response and peak glucose height were measured after rice consumption. A general downward trend in peak glucose over time was observed, suggesting that continued scOat Fiber intake may moderate glycemic responses. The effect was most evident at higher intake levels, with notable reductions at 20 g/day (Figure 2A). Moreover, this attenuation of glycemic excursions appeared to intensify with both dose and intervention duration, as reflected by the progressive decreases across T1, T2, and T3 in the 20 g/day group (Figure 2B). A significant reduction in peak glucose between interventions T0 and T3 was revealed in the 10 and 20 g/day groups (FDR-adjusted Student’s t test, *p*= 0.0227 and 0.0177; Table 3). Furthermore, when comparing peak glucose during rice challenge at intervention T3 vs rice challenge at intervention T0, a reduction was observed in 54% of participants from the 5 g/day group, 69% of participants from the 10 g/day group and 83% of participants from the 20 g/day group.

**Table 3.**
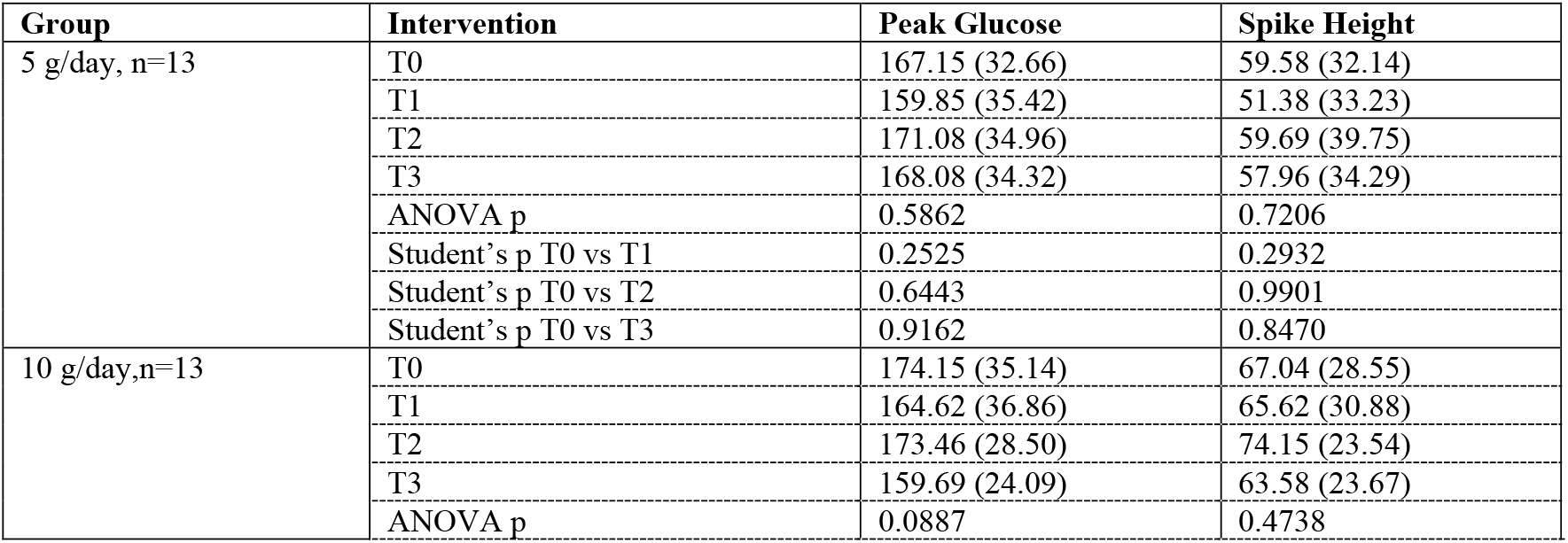
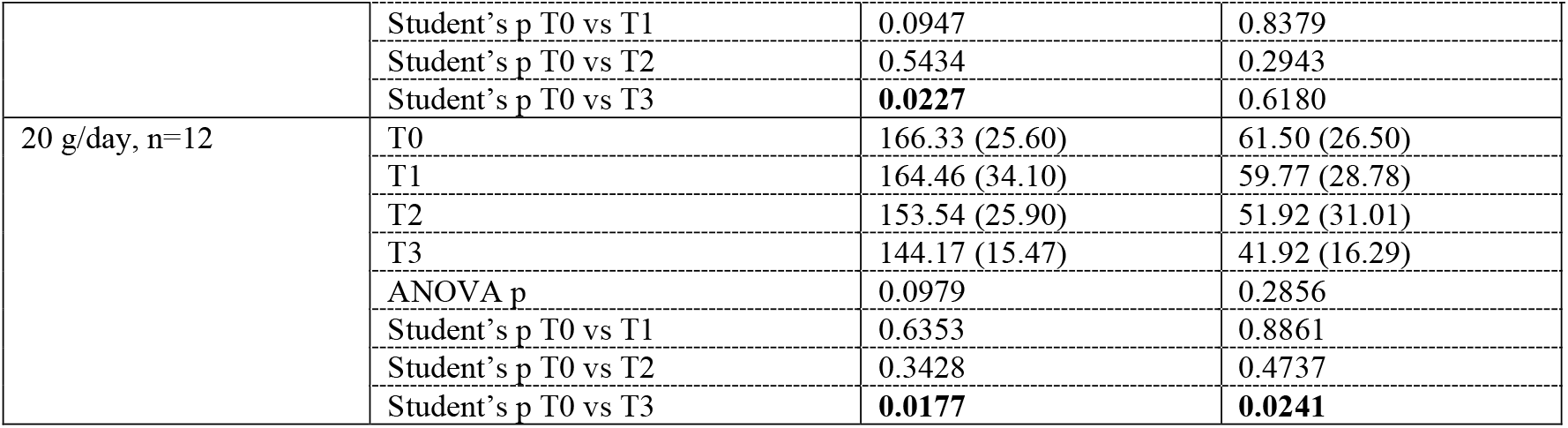
Peak glucose and glucose spike height during rice challenges. Values represent mean (SD) for each group receiving scOat Fiber at different doses. Statistical significance of changes at intervention period compared to Intervention T0 (ANOVA, FDR-adjusted Student’s p value) are reported.

**Figure 2.**
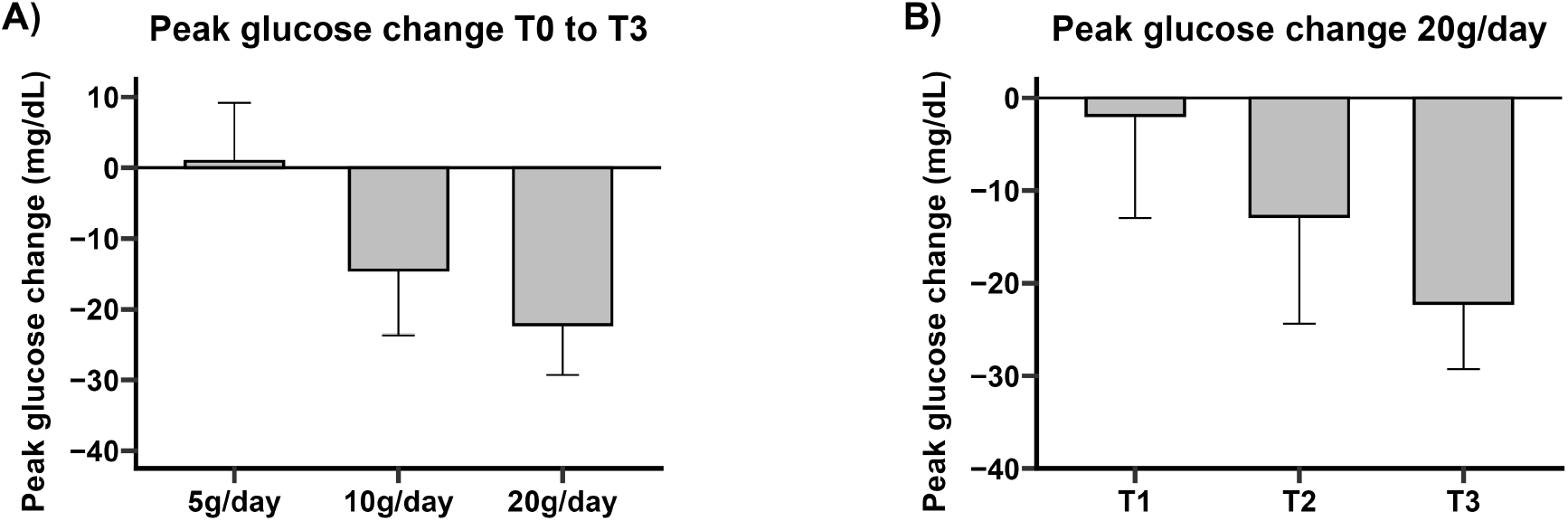
Effect of scOat Fiber on peak glucose during rice cereal challenges. A) Change of peak glucose during rice challenges performed at intervention T3 vs intervention T0 in all participants; B) Change in peak glucose during rice challenges performed at interventions T1, T2 and T3 in 20 g/day group. Error bars represent standard error of the mean.

Similar outcomes were seen for the glucose spike height. The glucose spike height showed a similar dose-dependent reduction, with the 20 g/day group demonstrating a significant decrease in glucose spike height between interventions T0 and T3 (FDR-adjusted Student’s t test, *p*= 0.024; Table 3).

Additionally, postprandial glycemic exposure was assessed using total AUC and iAUC during rice challenges, at intervention T1, intervention T2 and intervention T3, compared to the rice challenge at intervention T0. Total AUC and iAUC of each group generally trended towards lower values at interventions T2 and T3, although statistical significance was not found for any dose or at any intervention (ANOVA, Student’s t-test; Table 4).

**Table 4.**
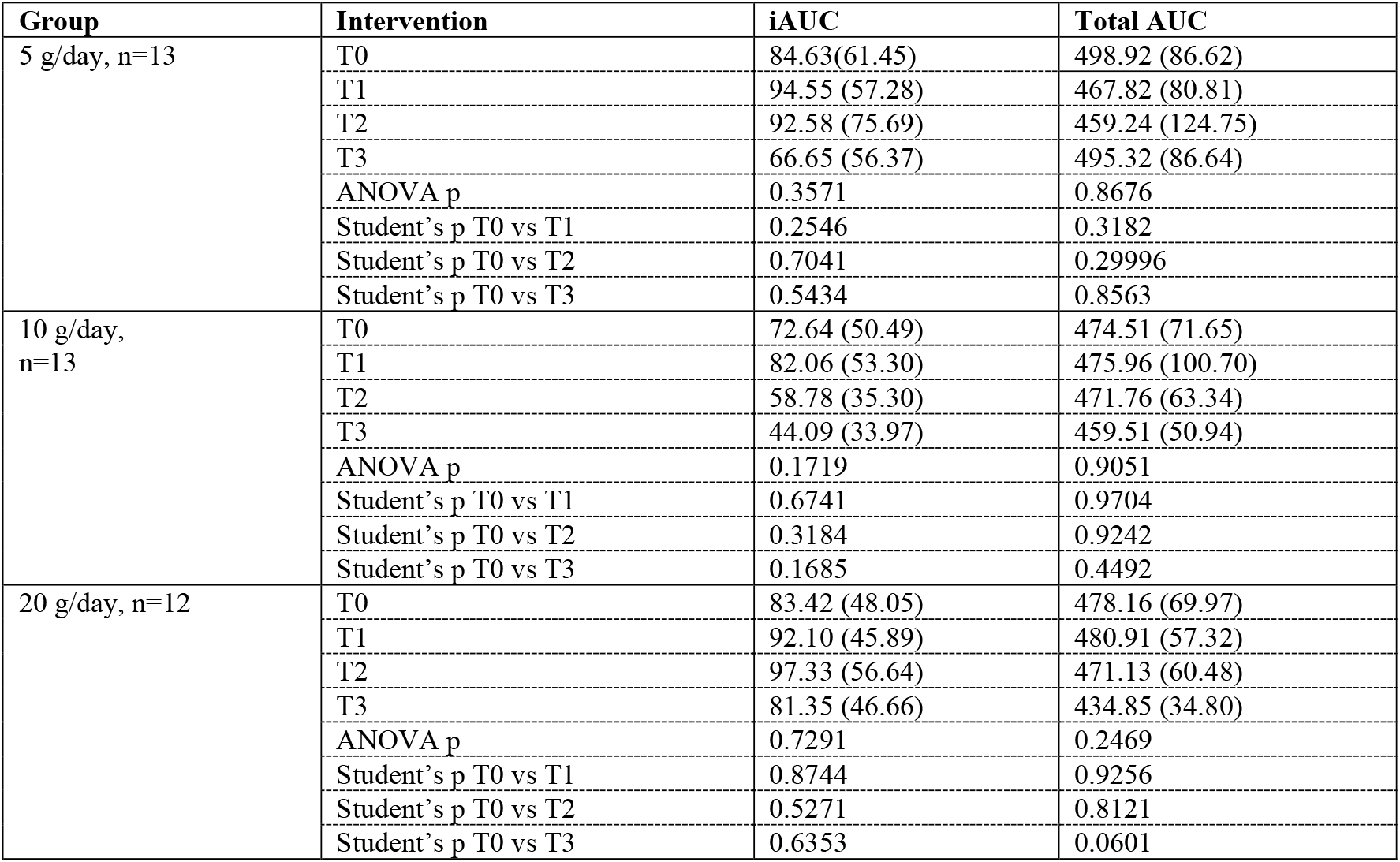
Incremental (iAUC) and total area under the curve (AUC) glucose responses during rice challenges. Values represent mean (SD) for each group receiving scOat Fiber at different doses. Statistical significance of changes at intervention period compared to Intervention T0 (ANOVA, FDR-adjusted Student’s p value) are reported.

| Group | Intervention | iAUC | Total AUC |
| --- | --- | --- | --- |
| 5 g/day, n=13 | T0 | 84.63(61.45) | 498.92 (86.62) |
|  | T1 | 94.55 (57.28) | 467.82 (80.81) |
|  | T2 | 92.58 (75.69) | 459.24 (124.75) |
|  | T3 | 66.65 (56.37) | 495.32 (86.64) |
|  | ANOVA p | 0.3571 | 0.8676 |
|  | Student's p T0 vs T1 | 0.2546 | 0.3182 |
|  | Student's p T0 vs T2 | 0.7041 | 0.29996 |
|  | Student's p T0 vs T3 | 0.5434 | 0.8563 |
| 10 g/day, n=13 | T0 | 72.64 (50.49) | 474.51 (71.65) |
|  | T1 | 82.06 (53.30) | 475.96 (100.70) |
|  | T2 | 58.78 (35.30) | 471.76 (63.34) |
|  | T3 | 44.09 (33.97) | 459.51 (50.94) |
|  | ANOVA p | 0.1719 | 0.9051 |
|  | Student's p T0 vs T1 | 0.6741 | 0.9704 |
|  | Student's p T0 vs T2 | 0.3184 | 0.9242 |
|  | Student's p T0 vs T3 | 0.1685 | 0.4492 |
| 20 g/day, n=12 | T0 | 83.42 (48.05) | 478.16 (69.97) |
|  | T1 | 92.10 (45.89) | 480.91 (57.32) |
|  | T2 | 97.33 (56.64) | 471.13 (60.48) |
|  | T3 | 81.35 (46.66) | 434.85 (34.80) |
|  | ANOVA p | 0.7291 | 0.2469 |
|  | Student's p T0 vs T1 | 0.8744 | 0.9256 |
|  | Student's p T0 vs T2 | 0.5271 | 0.8121 |
|  | Student's p T0 vs T3 | 0.6353 | 0.0601 |

Together, these findings suggest that scOat Fiber may exert a dose and time dependent effect on postprandial glucose regulation, with the strongest effects observed after two weeks at the 20 g/day dose.

### 3.5 Impact of scOat Fiber on glucose variability

To evaluate the impact of daily scOat Fiber consumption on glucose variability, metrics including glucose mean, Standard Deviation (SD), Coefficient of Variation (CV), mean amplitude of glucose excursion (MAGE), Time in Range (TIR), Ideal Time in Range (iTIR) and all excursions were calculated for the period prior to intervention T1, and the five days before intervention T2 and the five days before intervention T3. The analysis included only participants who had complete CGM data across all assessed time points (n=35).

A significant increase in TIR over time was observed in both the 10 g/day and 20 g/day dose groups (ANOVA, *p*= 0.0377, *p*= 0.0231). TIR increased significantly between interventions T0 and T3 in both 10 g/day and 20 g/day group (FDR-adjusted Student’s p value, *p*= 0.0248 and *p*= 0.0189, respectively). TIR also increased significantly between interventions T0 and T2 in the 20 g/day group (FDR-adjusted Student’s p value, *p*= 0.0393) (Table 5, Figure 3A). Additionally, iTIR increased significantly between interventions T0 and T3 in the 20 g/day dose group (FDR-adjusted Student’s p value, *p*= 0.0411) (Table 5). During the last five days before the end of the study and compared with baseline period (before intervention T1), an iTIR increase was observed in 69% of participants from 5 g/day group, 83% of participants from 10 g/day group and 90% of participants at 20 g/day group. Also, glucose SD decreased in both 10 g/day and 20 g/day groups (ANOVA, *p*= 0.0062 and *p*= 0.0005, indicating a reduction in overall day-to-day fluctuations. Both 10 g/day and 20 g/day groups also demonstrated a significant decline in SD when comparing interventions T0 and T3 (FDR-adjusted Student’s p value, *p*= 0.0080, *p*= 0.0003) (Table 5, Figure 3B). MAGE values were reduced in the 10 g/day group (ANOVA, *p*= 0.0446), reflecting attenuation of glucose excursions. These findings were further supported by a reduction in the total number of excursions outside of the target range in 20 g/day group (ANOVA, *p*= 0.0045). Significance in the reduction was found when comparing interventions T0 vs T2, and intervention T0 vs T3 (FDR-adjusted Student’s p value, *p*= 0.0133 and *p*= 0.0059) (Table 5). Non-significant trends toward reduction in glucose mean and CV was observed for participants receiving 10 g/day and 20 g/day doses.

**Table 5.** Glycemic control metrics. Glycemic control metrics calculated for the period prior to intervention T1 (Baseline) and for the final five days of product use before intervention T2 (Week 1) and before intervention T3 (Week 2). Values represent mean (SD), n=38. Bold values indicate significant time-trends (ANOVA, FDR-adjusted Student’s p value, *p*< 0.05).

|  | 5 g/day | 10 g/day | 20 g/day |
| --- | --- | --- | --- |
| **Glucose mean** |  |  |  |
| Baseline | 112.41 (10.11) | 110.22 (12.29) | 116.68 (11.97) |
| Week 1 | 113.59 (11.30) | 103.86 (9.74) | 108.96 (6.48) |
| Week 2 | 113.50 (10.95) | 102.45 (10.75) | 108.50 (4.06) |
| ANOVA $p$ | 0.1874 | 0.0684 | 0.0674 |
| Student's $p$ Baseline vs Week 1 | 0.6274 | 0.0801 | 0.0774 |
| Student's $p$ Baseline vs Week 2 | 0.6883 | 0.6661 | 0.0619 |
| **Glucose Standard Deviation** |  |  |  |
| Baseline | 19.65 (3.71) | 21.23 (4.47) | 21.43 (4.19) |
| Week 1 | 18.82 (4.76) | 20.04 (3.99) | 17.41 (2.30) |
| Week 2 | 18.27 (4.48) | 18.09 (3.98) | 16.96 (2.47) |
| ANOVA $p$ | 0.3792 | **0.0062** | **0.0005** |
| Student's $p$ Baseline vs Week 1 | 0.4851 | 0.8383 | 0.0736 |
| Student's $p$ Baseline vs Week 2 | 0.2779 | **0.0080** | **0.0003** |

| % Coefficient of Variation |  |  |  |
| --- | --- | --- | --- |
| Baseline | 17.48 (2.88) | 19.27 (3.59) | 18.36 (3.14) |
| Week 1 | 16.68 (4.32) | 19.22 (3.16) | 15.98 (1.91) |
| Week 2 | 16.20 (4.12) | 17.58 (3.19) | 15.65 (2.32) |
| ANOVA p | 0.3332 | 0.2408 | 0.0610 |
| Student's p Baseline vs Week 1 | 0.4482 | 0.9723 | 0.0791 |
| Student's p Baseline vs Week 2 | 0.2226 | 0.1730 | 0.0571 |
| MAGE |  |  |  |
| Baseline | 32.83 (7.65) | 34.29 (8.94) | 34.80 (9.33) |
| Week 1 | 33.27 (9.22) | 33.92 (7.34) | 31.49 (4.40) |
| Week 2 | 33.02 (9.66) | 30.79 (8.65) | 29.59 (5.52) |
| ANOVA p | 0.9394 | **0.0446** | 0.0787 |
| Student's p Baseline vs Week 1 | 0.8548 | 0.7906 | 0.1887 |
| Student's p Baseline vs Week 2 | 0.9431 | 0.0536 | 0.0526 |
| All Excursions |  |  |  |
| Baseline | 14.56 (2.63) | 16.24 (3.77) | 15.89 (2.79) |
| Week 1 | 14.07 (3.29) | 15.23 (3.43) | 13.05 (1.98) |
| Week 2 | 13.56 (3.15) | 13.89 (3.31) | 12.67 (2.01) |
| ANOVA p | 0.3825 | 0.1325 | **0.0045** |
| Student's p Baseline vs Week 1 | 0.5697 | 0.4570 | **0.0133** |
| Student's p Baseline vs Week 2 | 0.2727 | 0.0698 | **0.0059** |
| Time in Range (TIR, %) |  |  |  |
| Baseline | 88.82 (8.25) | 83.55 (11.08) | 84.93 (10.50) |
| Week 1 | 88.65 (7.32) | 90.39 (4.09) | 92.98 (4.17) |
| Week 2 | 90.30 (6.80) | 92.47 (3.68) | 94.55 (2.41) |
| ANOVA p | 0.4565 | **0.0377** | **0.0231** |
| Student's p Baseline vs Week 1 | 0.9347 | 0.0755 | **0.0393** |
| Student's p Baseline vs Week 2 | 0.4126 | **0.0248** | **0.0189** |
| Ideal Time in Range (iTIR%) |  |  |  |
| Baseline | 52.38 (19.49) | 51.48 (19.15) | 47.38 (23.02) |
| Week 1 | 49.24 (26.56) | 63.38 (22.28) | 60.80 (15.22) |
| Week 2 | 49.31 (26.88) | 66.29 (24.05) | 63.71 (9.39) |
| ANOVA p | 0.6513 | 0.0630 | 0.0734 |
| Student's p Baseline vs Week 1 | 0.5783 | 0.084 | 0.1306 |
| Student's p Baseline vs Week 2 | 0.5783 | 0.0530 | **0.0411** |

**Figure 3.**
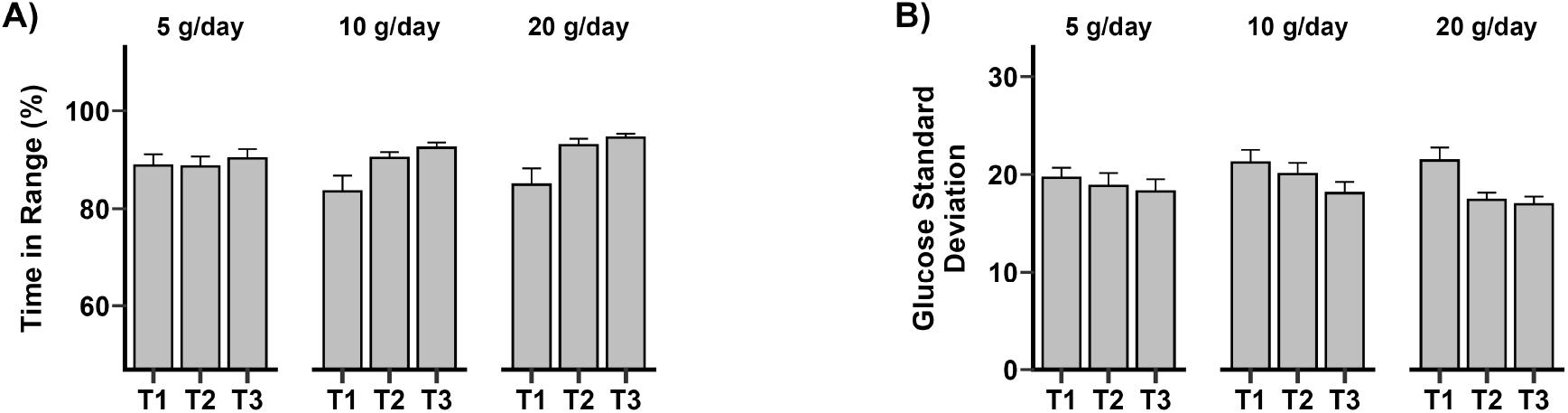
Effect of scOat Fiber on glycemic control metrics calculated for the period prior to intervention T1 (Baseline) and for the final five days of product use before intervention T2 (Week 1) and before intervention T3 (Week 2) in all participants. A) Change in TIR SD; B) Change in glucose SD. Error bars represent standard error of the mean.

The results indicate that, in addition to postprandial glucose control, scOat Fiber may have contributed to improved basal glycemia and overall improved glucose control in the 10 g/day and 20 g/day groups.

### 3.6. Exploratory outcomes: Mental health, appetite and sleep

To explore the mental health impact of scOat Fiber supplementation, participants completed weekly questionnaires incorporating selected questions from the validated GAD-7 and PHQ-9 assessment tools for mental health.

Due to the high proportion of participants reporting no mental health symptoms at intervention T0, a subgroup analysis was conducted on participants who reported at least mild symptoms (score ≥1 on a 0–4 scale) at intervention T0. The results from these symptomatic participants in all groups were pooled together. Domains such as anxiety, worry, irritability, anhedonia and life difficulty feeling, show significant reduction over time of symptoms (Friedman, *p*< 0.05). Significant decrease in anxiety, irritability and life difficulty was found between intervention T0 and interventions T2 (FDR-corrected Wilcoxon: anxiety, *p*= 0.0050, irritability; *p*= 0.0169, life difficulty, *p*= 0.0136). Anxiety, worrying, irritability, anhedonia, and life difficulty scores show a significant decrease at intervention T3, when compared to intervention 0 (FDR-corrected Wilcoxon, *p*= 0.0013, *p*= 0.0055, *p*= 0.0142, *p*= 0.0481, *p*= 0.0308, respectively) (Table 6).

**Table 6.**
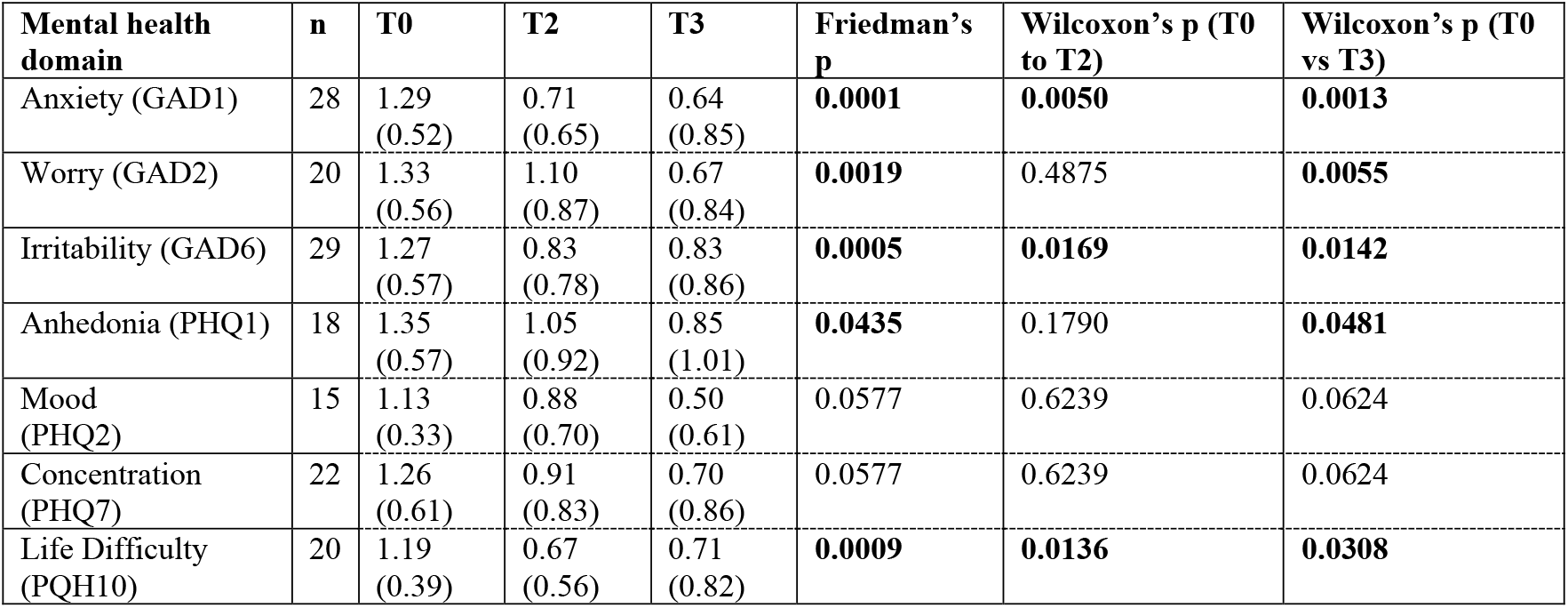
Impact of scOat Fiber on a subgroup of participants with mental health scores >1. Values represent mean (SD). Data corresponds to all doses pooled. Bold values indicate significant differences (p < 0.05).

In addition, participants completed a weekly questionnaire assessing appetite. Appetite scores remained stable over the 2-week intervention period across all groups (data not shown).

Daily questionnaire data were used to evaluate dietary deviations, change in perceived energy levels (including “morning slumps”), appetite and sleep duration. Daily sleep and appetite metrics did not vary significantly over time or between groups with fiber consumption for two weeks (Mann-Kendall, p>0.05). Rates of reporting morning slumps were infrequent, occurring in fewer 20% of participants in any group.

## 4 Discussion

This study evaluated the tolerability of scOat Fiber in healthy adults. This novel fiber is the product of controlled depolymerization of oat fiber, which significantly increases palatability and solubility of the original fiber. Oats fibers are well known to support healthy blood glucose levels and to have beneficial effects on postprandial blood glucose. However, incorporating enough for meaningful impact in diet requires consumption of large amounts of oat-based food. Approximately 20 g of scOat Fiber can deliver a fiber content equivalent to that found in four servings of oatmeal, offering an alternative approach to achieve similar diet intake and obtain metabolic and digestive benefits of oat fiber. From an organoleptic perspective, scOat Fiber has a broader range of food applications than oat fiber polysaccharide, making it a candidate for broad fiber supplementation. However, the gastrointestinal tolerance of scOat Fiber was unknown and therefore was evaluated. Three different amounts of fiber were tested for tolerability and safety. The results indicate that scOat Fiber was well tolerated and safe – even at the highest dose tested, 20 g/day. Across all the tested doses, participants reported no increase in gastrointestinal discomfort, with GSRS total scores remaining in the low range. Reported adverse effects assessed as possibly related to the scOat Fiber were mild, transient, and with no difference between doses.

Gastrointestinal symptoms, in fact, improved in the 5 g/day and 10 g/day groups despite the low level of symptoms at baseline, particularly in relation to overall symptoms (Total GSRS score) and the abdominal pain domain (GSRS abdominal pain). The improvement was seen after 1 week of intervention for the 5 g/day group and after two weeks for the 10 g/day group. Improvements were also seen in relation to the constipation domain after two weeks. Despite the improvement in constipation symptoms, no increase in diarrhea symptoms was provoked.

These results suggest that scOat Fiber may help address the chronic under-consumption of dietary fiber in the population by providing an alternative form of fiber that is well tolerated even at high doses. Unlike many other fibers, scOat Fiber did not provoke gastrointestinal intolerance and it was associated with improvement of some GI symptoms. This study counters the common consumer perception that all fibers cause digestive discomfort and highlights that fibers with specific structural characteristics can elicit diverse physiological responses. For example, inulin, which is commonly added to food and beverages, often provokes gastrointestinal intolerance at doses as low as 7.5 g/day, with studies reporting flatulence and bloating, and abdominal pain (Holscher et al., 2014; Ripoll et al., 2010). Studies with short chain fructans (FOS) also reported intolerance symptoms at doses above 5 g/day (Bonnema et al., 2010). Polydextrose has shown significant increase of bloating at doses of 6.25 g/day (Hull et al., 2012), and resistant dextrin can increase gastrointestinal symptoms at 7.5 g/day (Fastinger et al., 2008). The favorable tolerability of scOat Fiber may be related to its specific structural characteristics and abundance of shorter chain oligosaccharides. β-glucan, the primary component in oat fiber, is predominantly fermented by a narrower subset of gut microbiota compared to other fermentable fibers such as inulin or FOS. This selective fermentation profile may result in a more targeted SCFA production while limiting excessive microbial gas production. *In vivo* fermentation studies have shown that oat beta-glucan generates substantially lower gas volumes compared to highly fermentable fibers such as inulin, while still producing acetate, propionate and butyrate (Carlson et al., 2017).

Oat polysaccharides are associated with metabolic health benefits [13-15] and therefore the impact of scOat Fiber on metabolic outcomes was explored using continuous glucose monitoring. The results suggest that scOat Fiber may have a positive effect on metabolic health, particularly at higher daily amounts. Peak glucose after a rice challenge dropped significantly after 2 weeks of scOat Fiber consumption at amounts of 10 g/day and 20 g/day. A similar reduction was obtained for glucose spike height at 20 g/day. Furthermore, consumption of 20 g/day led to an average 13% decrease in peak glucose over 2 weeks, a magnitude of effect that aligns with reduction comparable to the magnitude of postprandial glycemic improvements considered physiologically beneficial in FDA evaluations of non-digestible carbohydrate intervention (Program, 2024). Contrary to peak glucose, no impact of scOat Fiber on the total exposure to glucose over the 4 hours after a rice challenge, as measured by AUC, was seen. A possible explanation is that scOat Fiber may lower the rate of glucose absorption, lowering the glucose peak but leaving the total amount of glucose taken up, unchanged. These findings suggest that the improvement in postprandial glucose metrics reflect more than just modulation of glucose absorption, previously described for scOat Fiber (Marcobal et al., 2024). A potential mechanism is that scOat Fiber may improve metabolic functioning over time, for example improving insulin sensitivity, allowing for a quicker initial response that lowers peaks but simultaneously reducing glucose troughs.

Peak glucose and AUC represent distinct but complementary aspects of postprandial glucose handling, providing an insight of how scOat Fiber modulates the acute glycemic response to meals. In contrast, parameters like TIR, SD, CV and MAGE reflect broader metabolic regulation, capturing effects beyond a single meal. A potential improvement in metabolic functioning is supported by the results of this study, obtained in relation to these parameters, which are increasingly recognized as risk factors for metabolic syndrome and cardiovascular events (Belli et al., 2023; Guo et al., 2022). The results suggest that scOat Fiber not only minimizes postprandial glucose excursions, reducing the amplitude of glucose spikes, but also may improve basal glycemia, a clinically meaningful effect for metabolic health.

While most studies emphasize beta-glucan viscosity as a key mechanism in glycemic control, this study demonstrates, for the first time, that cereal beta-glucans improve blood glucose outcomes independent of their viscosity. This study aligns with recent research which indicates that short chain or non-viscous fibers may offer glycemic benefits, especially when controlling for food dose. It has been shown that low molecular weight β-glucan performs similarly to high molecular weight β-glucan on metrics such as iAUC or peak glucose (Ames et al., 2021). Importantly, a study on evening intake of cereal β-glucan found improvements in the next day’s glycemia despite no clear dependence on viscosity, pointing to a possible gut microbiome mediated effect (Telle-Hansen et al., 2022). Production of SCFA by fiber fermentation may impact glucose dynamics as described before (Slavin, 2013), potentially improving glucose handling and insulin sensitivity.

In an exploratory analysis, participants with mild mental symptoms (average score in the baseline mood questionnaire of 1 or higher) reported significant improvement after two weeks of consuming scOat Fiber. Consistency of the changes across multiple symptom domains and the statistical significance of the comparisons suggest that scOat Fiber may support mental health and this potential positive effect is worth further study. While these findings were statistically significant and consistent, they must be interpreted as exploratory due to the absence of a control group, pooling of the participants and the small subgroup sample size. Nonetheless, they align with growing evidence in the literature linking fiber intake and mental health, thus future evaluations should be made with participants that present elevated symptom scores following expert recommendations (Dalile et al., 2025). Mechanistically, the link between fiber and mental health is supported by an extensive body of research. SCFAs produced in the gut as a product of fiber fermentation, are absorbed and can cross the blood brain barrier, reduce brain inflammation and affect neurotransmitters that regulate mood, such as serotonin, GABA and dopamine (Wolever et al., 2021). In addition, fibers can improve gut barrier function and lower levels of circulating toxins, which may reduce overall inflammation and the impact on gut-brain function (Kelly et al., 2015). Similar benefits have been seen with other fibers, such as psyllium, which has been linked to better mood, and inulin, which may improve both mood and cognitive function (Johnstone and Cohen Kadosh, 2025; Schmidt et al., 2015). There is also evidence that galacto-oligosaccharides (GOS) and resistant starch may offer improvements in overall mental health (Johnstone et al., 2021; Kadyan et al., 2024).

Several limitations of the current study should be acknowledged. The open label design and lack of a placebo control limit the ability to distinguish intervention effects from placebo effects, particularly for subjective outcomes such as mood and sleep. Further, potential confounding factors such as baseline fiber intake, exercise and sleep were not taken into account. Also, the intervention period was relatively short (14 days), and long-term benefits remain unknown. Further, while multiple glucose challenges were conducted, variability in participant adherence (i.e. exact rice intake timing, fasting) may have introduced noise into the CGM data. While stool samples were collected for microbiome and SCFA analysis, these results will be reported at a later time. Future publications integrating these data will help clarify the biological pathways through which scOat Fiber exerts its effects.

Although the current study did not include a placebo control, comparison between dose groups can partially compensate for the absence of a placebo control. Also, the strength and trends of the observed effects on gastrointestinal symptoms, glucose control and mental symptoms suggest that those effects may be different than a nonspecific placebo response. Future randomized controlled trials are needed to confirm these findings and to better understand dose response effects and underlying mechanisms. scOat Fiber is non-viscous, stable in food and beverage applications, and easy to incorporate into beverages, food and supplements, without compromising texture or palatability. Being able to incorporate this novel fiber as a food ingredient or supplement is a safe and scalable strategy to incorporate products that can support glycemic and mental well-being.

## 5 Conclusions

As many low tolerability fibers are added to foods or used as supplements, determination of tolerance to novel fibers is important. The findings from this study provide evidence that scOat Fiber, a short chain β-glucan, is safe and well tolerated, even at high daily intakes. This high tolerability allows food formulation with fiber dose levels that can beneficially impact consumers and may help addressing the current “fiber gap” in western diets. scOat Fiber shows functional properties relevant to gastrointestinal, metabolic and mental health. The results justify further investigation in populations with affected glucose regulation or participants with mild mental health symptoms. Given the global underconsumption of fiber and its implication in health, the development of tolerable and functional fibers represents a significant opportunity to improve public health.

## Data Availability

All data produced in the present study are available upon reasonable request to the authors

## 7 Author Contributions

Conceptualization AM. M., KM. N., BR. M., MJ. A.; Data Curation KM. N., BR. M, Writing-Original draft preparation: AM.M, Writing-Review & Editing: BR.M., KM. N., RA. D., MJ. A., Supervision AM.M., Project Administration AM. M., RA. D.

## 9 Acknowledgments

The authors thank the study participants and research team at People Science Inc. for conducting and supporting this work

## 10 Data Availability Statement

The data presented in this study are available on request due to privacy reasons.

