## Supplemental Information for "Short-Chain Oat Fiber Improves Gastrointestinal Tolerance and Regulates Glucose Metabolism: A Two-Week Open-Label Study in Healthy Adults"

### *Supplementary Material*

**Supplementary Table S1.** GSRS subcategories scores across the study period. Values represent mean (SD), for each group receiving scOat Fiber at different doses. Statistical significance of changes over time (Friedman test) and between intervention T0 and interventions T2 and T3 (Wilcoxon test) is reported as p-values. Significant results ($p<0.05$) are highlighted in bold.

| | **5 g/day (n=24)** | **10 g/day (n=19)** | **20 g/day (n=20)** |
|---|---|---|---|
| **Reflux** | | | |
| Intervention T0 | 1.3 (0.6) | 1.3 (0.5) | 1.3 (0.7) |
| Intervention T2 | 1.1 (0.4) | 1.3 (0.6) | 1.3 (0.7) |
| Intervention T3 | 1.1 (0.2) | 1.2 (0.3) | 1.5 (0.8) |
| Friedman p | 0.0862 | 0.2231 | 0.5529 |
| Wilcoxon p (T0 to T2) | 0.3020 | >0.9999 | 0.7518 |
| Wilcoxon p (T0 to T3) | 0.2382 | 0.4533 | 0.7518 |
| **Abdominal Pain** | | | |
| Intervention T0 | 1.8 (0.7) | 1.4 (0.5) | 1.6 (0.7) |
| Intervention T2 | 1.4 (0.4) | 1.3 (0.5) | 1.5 (0.6) |
| Intervention T3 | 1.4 (0.4) | 1.1 (0.2) | 1.8 (1.0) |
| Friedman p | **0.0005** | **0.0074** | 0.6278 |
| Wilcoxon p (T0 to T2) | **0.0012** | 0.1944 | 0.4768 |
| Wilcoxon p (T0 to T3) | **0.0122** | **0.0187** | >0.9999 |
| **Indigestion** | | | |
| Intervention T0 | 2.3 (0.7) | 1.9 (0.8) | 1.9 (0.8) |
| Intervention T2 | 1.9 (1.1) | 1.7 (0.6) | 1.9 (0.9) |
| Intervention T3 | 1.9 (1.0) | 1.5 (0.5) | 2.0 (1.1) |
| Friedman p | 0.0670 | 0.4395 | 0.8052 |
| Wilcoxon p (T0 to T2) | **0.0465** | 0.9336 | 0.6926 |
| Wilcoxon p (T0 to T3) | 0.0768 | 0.3593 | 0.8744 |
| **Diarrhea** | | | |
| Intervention T0 | 1.9 (1.1) | 1.2 (0.4) | 1.5 (0.8) |
| Intervention T2 | 1.4 (0.7) | 1.2 (0.3) | 1.6 (0.9) |
| Intervention T3 | 1.5 (1.0) | 1.0 (0.1) | 1.4 (0.6) |
| Friedman p | 0.0921 | 0.2263 | 0.4281 |
| Wilcoxon p (T0 to T2) | 0.1124 | 0.7456 | 0.4047 |
| Wilcoxon p (T0 to T3) | 0.3123 | 0.2561 | 0.5050 |
| **Constipation** | | | |
| Intervention T0 | 1.8 (1.0) | 1.8 (1.0) | 2.0 (1.2) |
| Intervention T2 | 1.6 (0.7) | 1.8 (1.1) | 1.6 (0.8) |
| Intervention T3 | 1.4 (0.7) | 1.5 (0.8) | 2.0 (1.5) |
| Friedman p | **0.0408** | **0.0204** | 0.0632 |
| Wilcoxon p (T0 to T2) | >0.9999 | 0.3173 | 0.1331 |
| Wilcoxon p (T0 to T3) | 0.0833 | **0.0244** | 0.1138 |

**Supplementary Table S2.** Adverse events

| AE Type | Total Count of AE Occurrences | Count of participants impacted | Impacted in 5 g/day group | Impacted in 10 g/day group | Impacted in 20 g/day group |
|---|---|---|---|---|---|
| Gas | 30 | 26 | 11 | 9 | 6 |
| Bloating | 17 | 15 | 7 | 4 | 4 |
| Constipation | 15 | 13 | 4 | 4 | 5 |
| Abdominal pain | 7 | 7 | 4 | 1 | 2 |
| Diarrhea | 5 | 5 | 4 | 0 | 1 |
| Dizziness | 2 | 2 | 1 | 1 | 0 |
| Borborygmus | 1 | 1 | 0 | 0 | 1 |
| Fatigue | 1 | 1 | 1 | 0 | 0 |
| Loose Stool | 1 | 1 | 0 | 1 | 0 |
| Nausea | 1 | 1 | 1 | 0 | 0 |
| Reflux | 1 | 1 | 0 | 1 | 0 |
